# FairCareNLP: An AI-Driven Patient Review Analyzer for Healthcare

**DOI:** 10.1101/2025.11.13.25340187

**Authors:** Sayyed Mohammad Pourya Momtaz Esfahani, Davey Seeman, Christoffer Dharma, Mohammad Noaeen, Shion Guha, Zahra Shakeri

## Abstract

**Objective:** To develop and evaluate an automatic patient review analyzer that applies advanced Natural Language Processing (NLP) and machine learning methods to improve the efficiency, fairness, and accuracy of healthcare feedback analysis.

**Materials and Methods:** We designed a multi-component pipeline incorporating sentiment analysis, key theme extraction, clinical Named Entity Recognition (NER), and fairness modules. Bias mitigation was addressed through the integration of three complementary approaches: adversarial debiasing, Hard Debiasing, and Iterative Null-space Projection (INLP). Multiple BERT-based models (DistilBERT, BioBERT, RoBERTa, BERT-base-uncased) were trained and evaluated under varying hyperparameters and fairness/adversarial loss configurations. Model performance was assessed using accuracy, F1, recall, precision, AUC, Equalized Odds (EOD), and Word Embedding Association Test (WEAT) metrics.

**Results:** Adversarial loss (*λ*_adv_ *>* 0) consistently decreased model performance across accuracy, F1, precision, and recall. In contrast, Hard Debiasing and INLP improved WEAT scores while preserving or enhancing other metrics, with INLP yielding the best overall performance. Specifically, INLP with fairness loss improved EOD by 14%, gender WEAT scores by 15%, and achieved slight gains for ethnicity and socioeconomic WEAT scores. The best model achieved accuracy of 0.856, F1 score of 0.812, recall of 0.798, AUC of 0.961, and precision of 0.829. The key theme analysis module identified 82% of expert-labeled themes, though 21% of patient comments lacked expert labels for valence or related attributes.

**Discussion:** Our results demonstrate the trade-offs between fairness and performance in bias mitigation strategies. While adversarial debiasing reduced predictive accuracy, INLP and Hard Debiasing improved fairness without significant degradation in task performance. Gender bias proved easier to mitigate than multi-categorical features such as ethnicity and income, underscoring the need for fairness techniques tailored to multi-class sensitive attributes.

**Conclusion:** This work presents a comprehensive NLP pipeline for patient feedback analysis that integrates multiple debiasing strategies, offering an important step toward equitable AI in healthcare. The approach enhances both the fairness and accuracy of insights drawn from unstructured patient reviews, thereby supporting more inclusive patient-centered care.

## 1 Introduction

Patient feedback is an invaluable resource for healthcare organizations striving to enhance care quality and patient outcomes. Studies have shown that healthcare facilities that actively gather and respond to patient feedback witness significant improvements in key performance indicators. A study using systematic patient feedback in an inpatient psychiatric facility found lower readmission rates compared to national benchmarks: 6.1% at 30 days, 9.5% at 60 days, and 16.4% at 180 days [1]. Furthermore, the World Health Organization (WHO) emphasizes that prioritizing patient-centered care, which heavily relies on understanding and acting upon patient feedback, leads to better health outcomes and higher levels of patient satisfaction [2]. Despite its importance, the vast amount of unstructured patient feedback, often collected through surveys, social media, and online reviews, presents significant challenges in analysis and actionable insight extraction.

The advent of advanced technologies, particularly in the context of Natural Language Processing (NLP) and Artificial Intelligence (AI), offers promising solutions to these challenges. AI-powered tools are increasingly recognized for their ability to process large datasets with enhanced speed and accuracy, far surpassing traditional manual methods [3]. For instance, AI-driven analysis has been shown to reduce data processing time, allowing healthcare providers to respond more promptly and effectively to patient needs [4, 5]. Additionally, NLP techniques enable the extraction of nuanced insights from unstructured text, such as identifying key themes and sentiments within patient reviews, which can be crucial for improving healthcare delivery [6].

However, while these technological advancements present substantial opportunities, they also introduce new challenges, particularly regarding the presence of biases in AI models. Biases, whether based on gender, ethnicity, or socioeconomic status, can significantly skew the results of NLP analyses, which can result in the underrepresentation of certain groups and the perpetuation of existing disparities in healthcare [7]. A recent study found that AI models with unmitigated biases were 40% more likely to misclassify sentiments from minority groups, underscoring the critical need for robust bias mitigation strategies [8]. Furthermore, research has shown that predictive models trained predominantly on White American data perform worse when applied to African American populations, which emphasizes the need for diverse training datasets [9]. Many studies fail to report crucial demographic information. In a systematic review of 164 machine learning studies using Electronic Health Record (EHR) data, 64% did not report race or ethnicity and 92% omitted any information on socioeconomic status [10]. This lack of demographic reporting extends to text-based diagnostic applications. In a systematic review of 78 studies using clinical text for diagnosis, only 35.9% provided race data, and among those, 57.1% described study populations that were majority White [11]. These biases can lead to misdiagnoses and a lack of generalization in minority populations [12]. Addressing these issues is essential for developing equitable healthcare solutions that can accurately analyze sentiments across diverse patient populations and support more personalized and effective treatments. This requires continuous monitoring of AI models for bias, regular updates with diverse datasets, and collaboration between AI developers and healthcare professionals to ensure the technology serves all patients equally.

Given these considerations, our work is driven by the imperative to develop an AI-powered patient review analyzer that not only enhances the efficiency and accuracy of patient feedback analysis but also ensures that the insights generated are unbiased and representative of all demographic groups. This approach aims to contribute to a more equitable healthcare system where patient feedback truly informs and improves care delivery.

Several studies have explored the application of NLP in the analysis of patient reviews, particularly focusing on sentiment analysis and key theme extraction. For example, Greaves et al. [13] demonstrated the effectiveness of sentiment analysis in monitoring patient satisfaction in real time, using social media data as a primary source. Similarly, Nawab et al. [14] leveraged NLP to extract meaningful insights from patient surveys, which identified specific areas requiring improvement in hospital services. Also, studies such as Yuan et al. [15] have applied sentiment and topic analysis to hospital experience comments using NLP techniques, while Feizollah et al.’s scoping review (2025) identified that among 52 studies applying NLP to unstructured patient feedback, sentiment analysis appeared in 32, topic modelling in 15, and text classification in 7. These studies highlight the potential of NLP in transforming unstructured patient feedback into actionable insights. However, they also reveal significant gaps, particularly in addressing biases that may skew analysis results. Research has also focused on extracting key themes from patient reviews using techniques such as Latent Dirichlet Allocation (LDA). While LDA can uncover recurring themes within patient feedback [16], it has limitations, such as treating documents as unordered sets of words and requiring a pre-specified number of topics. These constraints can obscure bias-related issues and misrepresent minority viewpoints, thereby perpetuating health disparities and compromising patient care.

Bias detection and mitigation have emerged as critical areas of concern in the application of AI in healthcare. Bolukbasi et al. [17] introduced methods for debiasing word embeddings to reduce gender and ethnic biases in NLP models. This seminal work sparked further research into algorithmic fairness in healthcare AI. For instance, Chen et al. [18] demonstrated that clinical notes contain implicit biases that can lead to disparities in machine learning predictions across racial groups.

Adversarial debiasing techniques have also been developed to remove protected attributes from model representations through a process in which a predictive model is trained in opposition to an adversary network that attempts to infer sensitive attributes, thus enhancing fairness in sentiment analysis [19]. In the healthcare domain, Zhang et al. [20] applied adversarial debiasing to mitigate gender and age biases in clinical outcome prediction models. Similarly, Pfohl et al. [21] proposed a multi-task learning approach to balance fairness and predictive performance in EHR-based risk models. More recently, FairEHR-CLP utilizes contrastive learning for fairness-aware clinical predictions across multimodal EHR data [22], and the FAME framework optimizes both performance and equity via fairness-aware multimodal embeddings in EHR predictive tasks [23]. Alongside adversarial debiasing, other advanced strategies such as Hard Debiasing [17, 24], which projects pre-trained embeddings onto a bias-neutral subspace, and Iterative Null-space Projection (INLP) [25], which iteratively removes protected attribute information from learned representations, have emerged as complementary post-processing approaches. Despite these advances, there is still a considerable gap in integrating comprehensive bias mitigation strategies into NLP pipelines for healthcare applications. Existing models often address specific biases (e.g., demographic parity in classification outputs [26]; debiasing word embeddings to reduce gender/occupation associations [17]; clinician-in-the-loop bias auditing focused on race or age groups [27]), remain limited in scope and fall short of providing a holistic solution that ensures equity across all demographic groups. Our study builds on these prior efforts by taking incremental steps toward more comprehensive approaches. Rajkomar et al. [28] highlighted the challenges of fairness in machine learning for healthcare, emphasizing the need for careful consideration of data collection, model development, and evaluation processes. Recent work by Gichoya et al. [29] revealed persistent racial biases in medical imaging AI, underscoring the urgency of developing more robust debiasing techniques. Additionally, Obermeyer et al. [30] uncovered significant racial bias in a widely used algorithm for predicting health needs, demonstrating the far-reaching consequences of unchecked AI bias in healthcare systems.

To address these challenges, researchers have proposed frameworks for comprehensive bias assessment and mitigation in healthcare AI. For example, Char et al. [31] outlined ethical principles for the development and deployment of AI in medicine, emphasizing the importance of transparency, accountability, and fairness. Similarly, Ghassemi et al. [32] provided a roadmap for developing equitable and trustworthy clinical AI systems, advocating for interdisciplinary collaboration and rigorous evaluation of fairness metrics. As the field progresses, there is a growing recognition of the need for intersectional approaches to bias mitigation, as highlighted by Buolamwini and Gebru [33] in their work on gender and skin-type bias in facial analysis algorithms. Translating these insights to healthcare AI remains an active area of research, with promising directions including federated learning for privacy-preserving and fair model training [34] and causal inference techniques for unbiased decision support systems [35].

Moreover, the literature suggests that self-selection bias in online reviews remains a significant challenge. This bias arises when the individuals who choose to leave reviews are not representative of the entire patient population, potentially leading to an overemphasis on extreme opinions [36]. Hu et al. [37] demonstrated that online reviews often follow a J-shaped distribution, with an overrepresentation of very positive and very negative reviews. In the healthcare context, this bias can skew perceptions of care quality and patient satisfaction. This issue is compounded by the fact that many NLP models have not been adequately trained on diverse datasets, which limits their ability to generalize across different patient demographics. As highlighted by Larrazabal et al. [38], AI models in healthcare often underperform for underrepresented groups, potentially exacerbating existing health disparities. This underscores the critical need for diverse and representative training data in healthcare AI applications.

Our work addresses these gaps by developing FairCareNLP, a comprehensive NLP pipeline that integrates multiple components to enable a holistic analysis of patient reviews. The primary objective of FairCareNLP is to provide an AI-powered tool capable of performing sentiment analysis, key theme extraction, clinical named entity recognition (NER), and bias detection and mitigation in a unified framework. This approach aligns with recent calls for more integrated and ethically-aware AI systems in healthcare, as advocated by Char et al. [31] and Ghassemi et al. [32]. This tool is designed to leverage fine-tuned BERT models such as DistilBERT, BioBERT, RoBERTa, and BERT-base-uncased, which are enhanced with advanced bias mitigation techniques to assess patient sentiment and satisfaction accurately. The use of these state-of-the-art language models builds upon the work of Lee et al. [39] on BioBERT and Liu et al. [40] on RoBERTa, adapting these powerful models to the specific challenges of healthcare text analysis.

A key component of our methodology is the application of three advanced bias mitigation techniques—Adversarial Debiasing, Hard Debiasing, and Iterative Null-space Projection (INLP). These represent distinct approaches applied at different stages of the model pipeline: adversarial debiasing is an in-processing method that trains the model to avoid encoding protected attributes through adversarial objectives; both Hard Debiasing and INLP are post-processing techniques—Hard Debiasing adjusts pre-trained embeddings via projection onto a neutral subspace, while INLP iteratively removes protected attribute information from learned representations through null-space projection—after model training but before inference [17, 24, 25]. By integrating all three strategies within a single NLP pipeline for patient review analysis, rather than applying them in isolation as prior studies have done, our work provides a more comprehensive and rigorous approach to mitigating bias. Together, these complementary methods enable a more comprehensive mitigation of bias throughout the training and inference pipeline, helping ensure our sentiment analysis remains equitable across patient demographics.

Another significant aspect of our work is the integration of a key theme prediction module, which employs the LLaMA language model to identify recurring themes and topics in patient reviews [41]. This module is particularly important for highlighting common concerns and areas for improvement in healthcare services, thereby providing actionable insights that can directly inform healthcare strategies.

The significance of our work lies in its potential to transform how healthcare organizations utilize patient feedback, making a substantial impact in the field of healthcare analytics. By providing a robust and comprehensive tool for analyzing patient reviews, we aim to significantly enhance the accuracy and fairness of insights generated from this data. Our approach not only improves the efficiency of feedback analysis but also ensures that all patient voices are equitably represented in the outcomes, contributing to the development of more responsive and inclusive healthcare systems. This aligns with the vision outlined by Rajkomar et al. [28] for ensuring fairness in machine learning to advance health equity. By addressing the challenges of bias, representativeness, and comprehensive analysis in patient feedback, our research contributes to the broader goal of leveraging AI to enhance healthcare quality and equity, as emphasized by recent studies such as Gichoya et al. [29] and Obermeyer et al. [30]. Ultimately, our work responds to the growing recognition that equitable and actionable insights are essential for improving patient-centered care and healthcare outcomes in an increasingly diverse and complex healthcare landscape.

## 2 Materials and methods

### 2.1 Dataset

This research utilizes a dataset obtained from patient surveys administered by the National Research Corporation (NRC), a US-based organization located in Ontario, Canada. These surveys encompass both quantitative metrics and open-ended responses regarding hospital experiences, offering a rich source of self-reported patient feedback. The Investigative Journalism Bureau (IJB), affiliated with the University of Toronto’s Dalla Lana School of Public Health, acquired this deidentified data spanning five years through freedom-of-information requests. The resulting compilation includes more than 120,000 anonymized patient reviews from 45 hospitals across Ontario, covering the period from 2015 to 2022. To ensure privacy, all names and identifiable data were replaced with X strings in the narrative text; no information can be traced back to participants. Ontario hospitals and health networks distributed these surveys, which captured various aspects such as hospital attributes (e.g., name, type, units), patient experience (e.g., patient reviews, visit date, sentiment valence), and key themes affecting patient satisfaction (e.g., respect, transportation, access, coordination). The dataset has been annotated to allow for the possibility of multiple key themes being associated with each patient review. Analysis were completed from January to August 2024.

To address the lack of demographic information in our deidentified dataset, we leveraged multiple authoritative sources to construct a representative profile of the population served by the hospitals in our study. We synthesized data from *Statistics Canada*, hospital stays data from the *Canadian Institute for Health Information*, and health care experience survey data from the *Ontario Ministry of Health* to extract the distributions of males and females, different age groups, ethnicities, and socioeconomic factors including Income/Poverty Level, Employment Status, Access to Transportation, and Educational Attainment for the cities in which hospitals were located. Using these distributions, we generated synthetic demographic attributes (e.g., gender, age group, ethnicity) for the reviews at each hospital by randomly assigning labels to comments in proportion to the real-world distributions. For instance, if a hospital’s patient population was 60% male and 40% female, we randomly labelled the hospital’s reviews using these probabilities. We then used a combination of socioeconomic distributions and considerations relative to gender, age, and ethnicity to generate socioeconomic data. For example, we adjusted the distribution for ages 65+ to reflect that they are mostly retired or not in the labor force, and for the 0-14 age group, we considered that they are mostly not in the labor force. Based on age groups or ethnicities, we applied some changes to the distributions and generated socioeconomic data, which gave us our model training dataset. Importantly, these generated demographic attributes were not used for training the sentiment, theme extraction, or NER models, nor for the operation of the bias mitigation techniques themselves. They were incorporated solely to enable the calculation of fairness metrics across demographic groups, ensuring that the evaluation of model outputs could be balanced against realistic population distributions. Because they were assigned independently of the review text, they did not introduce new patterns into the training data or influence the learned language representations. We also used the same method with modified distributions from other cities and countries to generate a mock demographic and socioeconomic dataset. This auxiliary dataset did not include any new or fabricated review text; instead, it provided a set of demographic and socioeconomic attributes constructed from realistic external distributions. Its sole purpose was to act as a comparison group with a different population distribution, simulating how an online reviewing population might differ from the institutional survey population. This allowed us to test the performance of our post-processing technique for mitigating self-selection bias in the propensity scoring module.

### 2.2 Sentiment Analysis Module

The primary predictive task in this module is sentiment analysis, i.e., classifying patient reviews by sentiment polarity (positive, negative, or neutral). Because demographic biases in model representations can distort sentiment predictions, this section details the debiasing strategies (INLP, Hard Debiasing, and Adversarial Debiasing) integrated into the sentiment analysis pipeline.

#### 2.2.1 Data Preprocessing

The preprocessing pipeline involves data cleaning by removing null values, duplicates, and irrelevant characters from the comments, standardizing visit dates by manually extracting various date formats and converting them to a consistent JavaScript Date type format, and correcting sentiment valence and key themes using the TextBlob library in Python to correct sentiment valence strings and key themes for consistent spelling. Large language models (LLMs) were used to map hospital units and types to refined categories, and encoding labels and sensitive features involved label encoding for sentiment labels and one-hot encoding for gender, ethnicity, and income. We tokenized comments using various BERT base models DistilBERT, BioBERT, RoBERTa, bert-base-uncased and tried to compare their performance, chunking long comments to ensure comments do not exceed the model’s maximum sequence length of 512 tokens, and padding shorter comments to maintain uniform input size.

#### 2.2.2 Tokenization and Chunking

To handle long comments effectively, we implemented a flagging and splitting process. This process is crucial for both model input and evaluation, as it ensures that all reviews, regardless of length, can be processed efficiently. Long reviews are flagged and split into manageable chunks, while shorter reviews are processed as-is. The algorithm for this process is outlined below:

**Fig 1.**
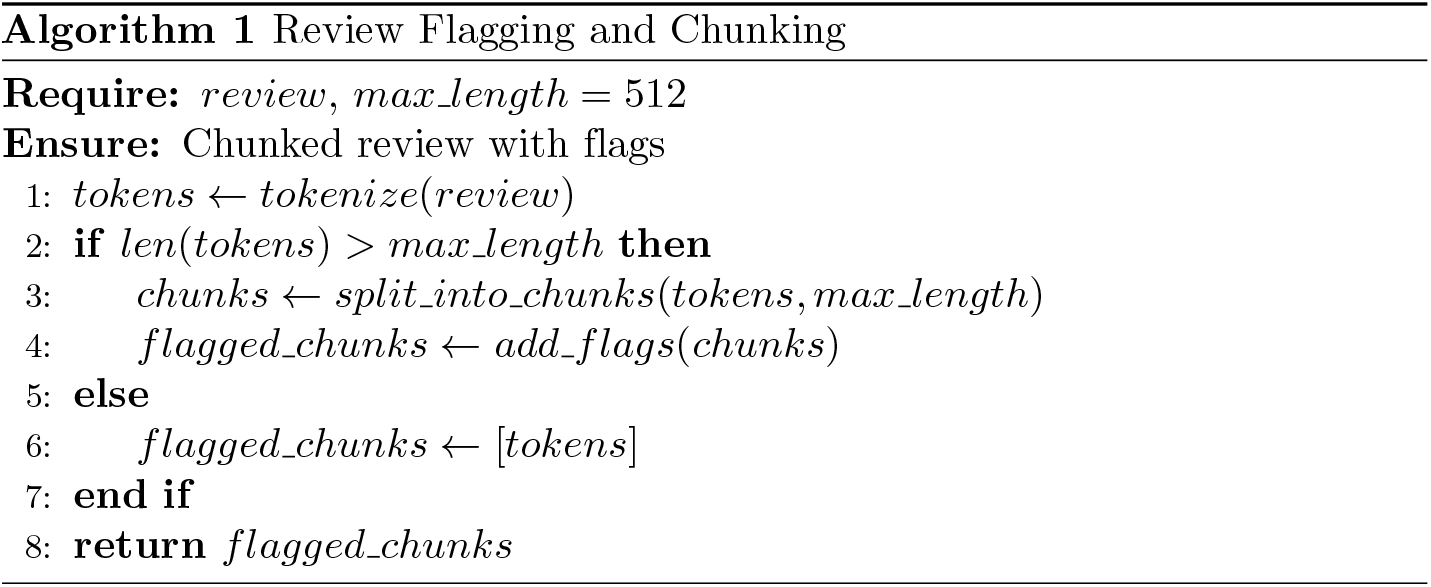
Pseudocode describing the review flagging and chunking process used in data preprocessing. Reviews exceeding the model’s maximum token limit (512) are divided into smaller segments, each tagged with positional flags to preserve context for subsequent analysis.

#### 2.2.3 Model Architecture

We used four different BERT base models DistilBERT, BioBERT, RoBERTa,bert-base-uncased for sentiment analysis, comparing their performance. The model architecture was modified to incorporate bias mitigation techniques. The last classifier layer was adjusted to match the number of sentiment labels in our dataset, and an adversarial classifier was added to predict protected attributes (gender, ethnicity) from the model’s internal representations. A gradient reversal layer was implemented between the main model and the adversarial classifier.

#### 2.2.4 Bias Mitigation Techniques

In our research, we explore and implement three powerful bias mitigation techniques to enhance the fairness of our language model: Iterative Null-space Projection (INLP), Hard Debiasing, and Adversarial Debiasing. Each method approaches the problem of bias from a different angle, providing a comprehensive strategy for mitigating various forms of bias in natural language processing models.

**Iterative Null-space Projection (INLP)** is a debiasing technique designed to systematically remove bias from the learned representations in a language model. The key idea behind INLP is to iteratively identify and project out the subspace where the bias manifests itself. This method ensures that the final embeddings do not encode information about the protected attributes, thus mitigating bias in downstream tasks.

The INLP method begins by identifying the bias direction, which is a vector in the embedding space that captures the bias. Mathematically, given an embedding matrix **E** ∈ ℝ^*d×n*^, where *d* is the dimensionality of the embeddings and *n* is the number of words, and a bias direction **v** ∈ ℝ^*d*^, the projection of an embedding **e**_*i*_ onto this bias direction is computed as:

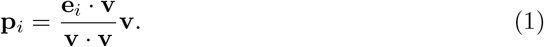

The debiased embedding is then obtained by subtracting this projection from the original embedding:

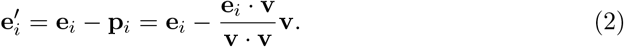

INLP extends this idea by iteratively projecting the embeddings onto the null space of the identified bias directions. After each iteration, a new bias direction is identified using a classifier, and the embeddings are further refined. The iterative update rule is given by:

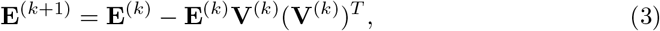

where **V**^(*k*)^ represents the matrix of bias directions identified in the *k*-th iteration. By repeating this process, INLP effectively removes the bias, making it difficult for the model to rely on protected attributes in its predictions [25]. In our implementation, we first initialize a BERT model without debiasing. We then use this model to compute the bias directions by analyzing the embeddings of word pairs that are known to exhibit bias. These directions are computed using Principal Component Analysis (PCA) on the difference vectors between such pairs. The resulting bias directions are then integrated into the BERT model, which is reinitialized with an INLP debiasing layer. This layer systematically removes bias from the embeddings during training.

**Hard Debiasing** is another powerful technique for mitigating biases in word embeddings. This method involves two primary steps: (1) neutralization and (2) equalization. The goal is to ensure that the embeddings of words associated with biased pairs (e.g., ‘he’ and ‘she’) are equidistant from a neutral point in the embedding space. The process begins by identifying the bias subspace using PCA on a set of biased word pairs. The top principal component(s) capture the bias direction. Given an embedding **e**_*i*_ and the bias subspace **B**, the neutralization step projects the embedding onto the subspace orthogonal to **B**:

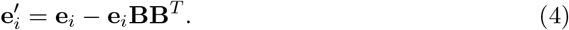

In the equalization step, pairs of words that should be equidistant (e.g., ‘he’ and ‘she’) are adjusted to ensure that their embeddings are equidistant from the mean of the pair. For a word pair (**e**_1_, **e**_2_), the mean embedding **e**_eq_ is calculated as:

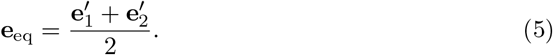

The equalized embeddings are then given by:

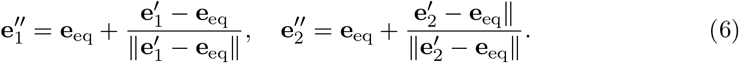

This process ensures that the embeddings do not favor one side of the bias spectrum over the other, thereby mitigating bias in the model’s predictions [17]. In our work, we compute the bias directions for gender, ethnicity, and income-related word pairs as described above. These directions are then used in a Hard Debiasing layer integrated into our BERT model. During training, this layer continuously adjusts the embeddings to reduce bias according to the principles of neutralization and equalization, resulting in more balanced representations across different demographic groups.

The integration of INLP and Hard Debiasing into our BERT-based sequence classification model is achieved through the inclusion of specialized debiasing layers. After computing the bias directions, we reinitialize the BERT model with these layers, which operate on the embeddings during both the forward pass and the backpropagation process. By doing so, we ensure that the learned representations are less likely to encode information related to protected attributes, thus enhancing the fairness of the model in real-world applications.

**Adversarial Debiasing** is a novel approach that leverages the power of adversarial learning to remove protected attributes from the model’s internal representations. In practice, this method trains two models simultaneously: the primary model performs the main prediction task (e.g., classifying review sentiment), while an adversarial network tries to predict sensitive attributes such as gender or ethnicity from the same internal representations. A gradient reversal layer ensures that as the adversary becomes better at detecting these attributes, the main model is pushed to learn representations where such attributes cannot be easily inferred. The result is that the model focuses on task-relevant features while minimizing dependence on protected demographic information, thereby improving fairness across groups [20, 42]. This function combines the primary task loss (sentiment analysis), fairness loss, and adversarial loss as follows:

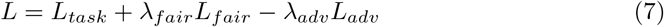

where *L*_*task*_ is the cross-entropy loss for sentiment classification, *L*_*fair*_ is the fairness loss, *L*_*adv*_ is the adversarial loss, and *λ*_*fair*_ and *λ*_*adv*_ are hyperparameters controlling the strength of fairness and adversarial components. Our custom loss function computes the loss for each chunk and adjusts it based on the number of words in chunks to handle long comments effectively.

To ensure fairness in our model’s predictions across different demographic groups, we incorporate a **fairness loss component** based on the principles of demographic parity and equalized odds. This approach aims to minimize disparities in the model’s performance among various sensitive features. The fairness loss is calculated by measuring the differences in true positive rates (TPR) and false positive rates (FPR) between groups. For a binary classifier *Ŷ* and a sensitive attribute *A*, demographic parity seeks to achieve:

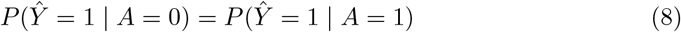

Similarly, equalized odds aims to equalize both TPR and FPR across groups:

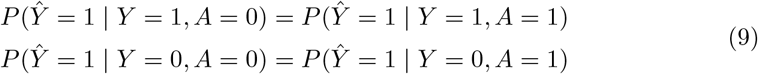

where *Y* represents the true label. To compute the fairness loss, we first calculate the TPR and FPR for each group *g*:

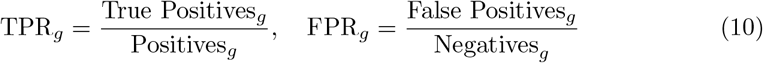

We then compute the absolute differences in TPR and FPR between groups:

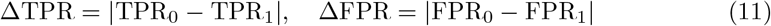

These differences are averaged across all labels *L*:

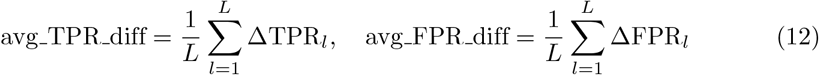

Finally, the fairness loss is calculated as the average of these differences:

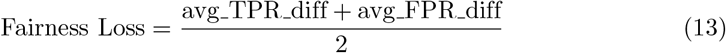

By incorporating this fairness loss into our overall loss function, we encourage the model to produce predictions that are more consistent across different demographic groups, thereby enhancing the overall fairness of our sentiment analysis system. The combination of INLP, Hard Debiasing, and Adversarial Debiasing provides a comprehensive approach to mitigating various forms of bias in our language model. Each technique addresses bias from a different perspective, allowing us to create a more robust and fair model for real-world applications.

#### 2.2.5 Training Process

The training process for our bias-mitigated sentiment analysis model incorporates several advanced techniques to optimize both performance and fairness. Specifically, we use a custom loss function that combines three elements: (1) the standard classification loss for predicting sentiment labels, (2) a fairness loss that encourages the model to perform equally well across demographic groups, and (3) an adversarial loss that trains the model to make it difficult for a secondary network to infer protected attributes such as gender or ethnicity. This comprehensive approach ensures that the model learns to perform sentiment analysis accurately while mitigating biases. To prevent overfitting—where a model performs well on training data but poorly on unseen data—we employ early stopping by monitoring the validation loss. Training is halted when the validation loss stops improving for a predefined number of epochs, helping to achieve the best generalization performance. Moreover, we apply dropout regularization with a rate of 0.1 in the BERT models. In this method, some of the connections in the neural network are randomly dropped during training, which forces the model to learn more robust patterns rather than memorizing noise from the training set.

For optimization, we use the `AdamW` optimizer, an extension of `Adam` that implements weight decay for additional regularization. The weight decay rate is set to 1e-5, providing a balance between model complexity and generalization ability. During training, we continuously apply projective debiasing to word embeddings. This process involves projecting the embeddings onto a subspace orthogonal to the identified bias directions, ensuring that the model learns fair representations throughout the training process. The model parameters are updated based on the gradients of the combined loss function, which includes the task loss, fairness loss, and adversarial loss components. This multi-faceted approach allows us to simultaneously optimize for performance and fairness.

#### 2.2.6 Evaluation

To comprehensively assess the effectiveness of our bias mitigation techniques, we employ a multi-faceted evaluation approach. The Word Embedding Association Test (WEAT) is a commonly used metric in bias evaluation literature; it measures the degree of association between sets of target words and attribute words in embedding space by comparing cosine similarities and using a permutation test to assess effect size and significance [43]. Positive WEAT effect sizes indicate stronger stereotypical associations, while values closer to zero suggest reduced bias. We also evaluate the model’s fairness using the equalized odds disparity metric, which measures the difference in true positive and false positive rates across protected groups. This metric helps us quantify the model’s consistency in performance across different demographic categories.

For performance evaluation, we use standard metrics such as accuracy, F1 score, precision, and recall. These metrics provide a comprehensive view of the model’s ability to correctly classify sentiments across various scenarios. To handle long comments that were split into chunks during processing, we employ a special evaluation procedure. We first reassemble the chunks using the flags added during the chunking process. For each reassembled review, we aggregate the probabilities of all its chunks. The final prediction for the review is computed as the weighted average of class probabilities across all chunks, with weights proportional to chunk lengths. We then calculate the evaluation metrics on these reassembled reviews, treating the aggregated prediction for all chunks of a review as a single prediction for that review. This approach ensures that our evaluation accurately reflects the model’s performance on complete reviews, even when they were processed in chunks.

### 2.3 Key Theme Analysis Module

After processing the review and extracting sentiments, we have our key theme analysis module. For key theme analysis, we engineered a prompt and wrapped the reviews with that prompt, using LLaMA3 70B to identify the key themes and the corresponding sections in the reviews for those key themes. We used the following prompt:

~~~
prompt = (“You are analyzing a patient review to identify key themes or areas
discussed in the text. “
“Key themes are specific topics, concerns, or aspects of the healthcare experience that the patient “
“Analyze the following patient review and identify all key themes from this list:
“
“mentions or talks about in their review.*\*n*\*n”
“Instructions:*\*n”
f”*{*‘, ‘.join(KEY THEMES)*}.\*n*\*n”
“-Identify themes that represent topics, concerns, or areas explicitly mentioned
or discussed in the review*\*n”
“-Match themes based on the content and context of what the patient is
“-A single review may contain multiple themes*\*n”
“-If no theme from the list matches the content, use ‘unknown’*\*n”
describing*\*n”
“-For each identified theme, provide a brief description explaining why this
theme applies*\*n*\*n”
“Respond with a JSON object containing a list of identified themes in the format
f”Patient Review:*\*n*{*patient review*}\*n*\*n”
“*{\*n”
below:*\*n”
“ *\*”themes*\*”: [*\*n”
“ *{\*n”
“ *\*”description*\*”: *\*”*\*”*\*n”
“ *\*”theme*\*”: *\*”*\*”,*\*n”
“]*\*n”
“ *}\*n”
“*}*”)
~~~

### 2.4 NER Module

After processing the review and extracting sentiment and key themes, we use the Bio-Epidemiology-NER model to tokenize, lemmatize, and extract entities such as disorders, chemicals, and drugs from patient reviews. The following is an example:

~~~
Review: “The nurse gave me acetaminophen for my migraine, which helped
with the pain but made me feel nauseous.”
Extracted Entities: Disorders: migraine, pain, nauseous Chemicals/Drugs:
acetaminophen
~~~

### 2.5 Propensity Scoring Module

One bias we anticipate within our online review system is selection bias, where certain groups of reviewers may be more likely to leave hospital reviews than others. A notable example of this bias is what is called the J-shaped curve bias, where strongly positive and strongly negative reviewers are more likely to leave reviews than those with neutral experiences. To address this potential bias, we developed a propensity scoring method to resample online respondents to match the distribution of in-person respondents across variables that could introduce selection bias, such as overall hospital experience. This approach ensures that reviewers who were less likely to respond are better represented in the resampled population, leading to a more balanced presence in our key themes classification model.

The variables considered, each potentially contributing to selection bias, were the following: overall hospital experience (valence), gender, age, ethnicity, hospital unit category, income, employment status, access to transportation, and educational attainment. We assumed that in-person reviews have less selection bias than online reviews, so controlling the variables to match the in-person distribution significantly reduces selection bias.

#### 2.5.1 Propensity Scoring Computation

Propensity scores in our context refer to the probability *P* (*T* = 1 | *X*) of a given reviewer belonging to the group of online respondents as opposed to in-person respondents based on the set of variables. Our goal was to balance the distribution of these variables to address selection bias.

In order to test the propensity scoring method, we used the synthetic demographic fields described in the Dataset subsection to construct a mock online cohort that approximated the characteristics of online reviewers. These fields did not include review text but were constructed to simulate how online reviewers might differ demographically from hospital-based respondents. We then trained a logistic regression model to predict the propensity scores *P* (*T* = 1 | *X*) from the synthetic online cohort and the survey data. The in-person dataset was drawn from the IJB dataset, which represent institutionally collected feedback. Although the dataset contains real review text, it does not specify the exact submission mode for each individual comment (e.g., paper, phone, or digital survey). We removed rows where the unit data was unknown. The model achieved an accuracy of 0.727 and an F1 score of 0.805, with the largest propensity score being 0.993 and the smallest as 0.084. The relevant dummy classifier achieved an accuracy of 0.647 and an F1 score of 0.786. A neural network with two hidden layers of 64 and 32 neurons achieved an F1 score of 0.804 and an accuracy of 0.728, similar to the logistic regression model’s performance. We chose to continue with the logistic regression model instead of the neural network for its simplicity and interpretability. Figure 2 displays the difference in propensity scores between online and in-person respondents.

**Fig 2.**
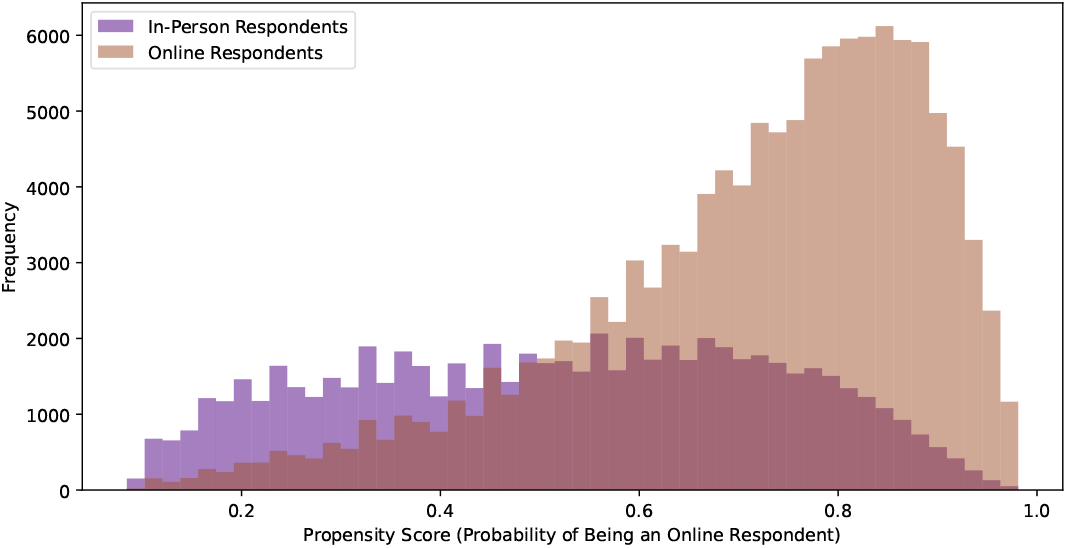
Propensity Score Distributions: Online vs. In-Person Respondents. This figure illustrates the propensity score distributions of online and in-person survey respondents, providing insights into the likelihood of participants choosing each survey method based on their characteristics.

#### 2.5.2 Propensity Scoring Strategies

The three primary propensity scoring techniques are propensity score matching, inverse propensity weighting (IPW), and propensity score stratification. We considered each of these methods and ultimately settled on propensity score stratification.

Propensity score matching involves matching each online reviewer to an in-person reviewer with the most similar propensity score. However, this approach may lead to many online reviews being matched to the same in-person review, potentially resulting in an overemphasis on these in-person reviews and causing biased classification.

Both IPW and propensity score stratification involve weighted resampling from the original population. In IPW, the (pre-normalized) resampling weight *w*_*i*_ for a respondent *i* is defined as follows:

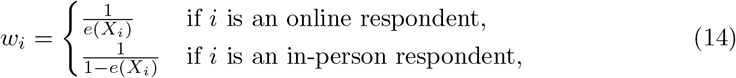

where *e*(*X*_*i*_) represents the propensity score of respondent *i*. This weighting ensures that after resampling, the groups resemble each other in terms of propensity scores and underlying variable distributions.

Propensity score stratification, on the other hand, involves resampling only from the treatment group (online distribution) and computing weights based on the relative representation of in-person to online respondents across propensity scores. The dataset is split into several strata (10 in our case) based on similar propensity scores. The relative representation *r*_*s*_ of in-person to online respondents within each stratum *s* is given by:

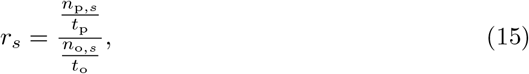

where *n*_p,*s*_ is the number of in-person reviewers within stratum *s, t*_p_ is the total number of in-person reviewers, *n*_o,*s*_ is the number of online reviewers within *s*, and *t*_o_ is the total number of online reviewers. The resampling weights *w*_*i*_ are then the normalized relative representations *r*_*s*_ for each respondent’s stratum.

Equation 14 defines the weights used in IPW, which ensures balanced representation between online and in-person respondents. Equation 15 is crucial in propensity score stratification, where it calculates the relative representation needed to adjust for the differences in the distribution of propensity scores between the two groups. This stratified approach allows for more nuanced control over the balance between treatment and control groups, ultimately leading to more accurate and less biased results.

### 2.6 System Integration and Workflow

Although each module was developed and evaluated independently, the overall design of the FairCareNLP pipeline envisions a future integrated framework for automated patient feedback analysis. In this conceptual system, patient reviews are first processed through the sentiment analysis, key theme extraction, and NER modules. The resulting outputs (sentiment labels, thematic categories, and extracted clinical entities) would then be aggregated within a hypothetical Intelligent Hospital System, an online software platform intended to facilitate continuous learning and healthcare quality monitoring. This platform would serve as a centralized analytics layer, storing intermediate outputs from all modules and generating dashboards for healthcare administrators. Before these aggregated data are visualized, the propensity scoring and distribution normalization module adjusts for demographic or self-selection biases, ensuring that reported insights more accurately reflect the overall patient population. While this integrated system is conceptual and not implemented in the present study, it illustrates the intended interoperability of the modules and the potential for real-world deployment within hospital analytics infrastructures. The overall workflow of this envisioned system is illustrated in figure 3.

**Fig 3.**
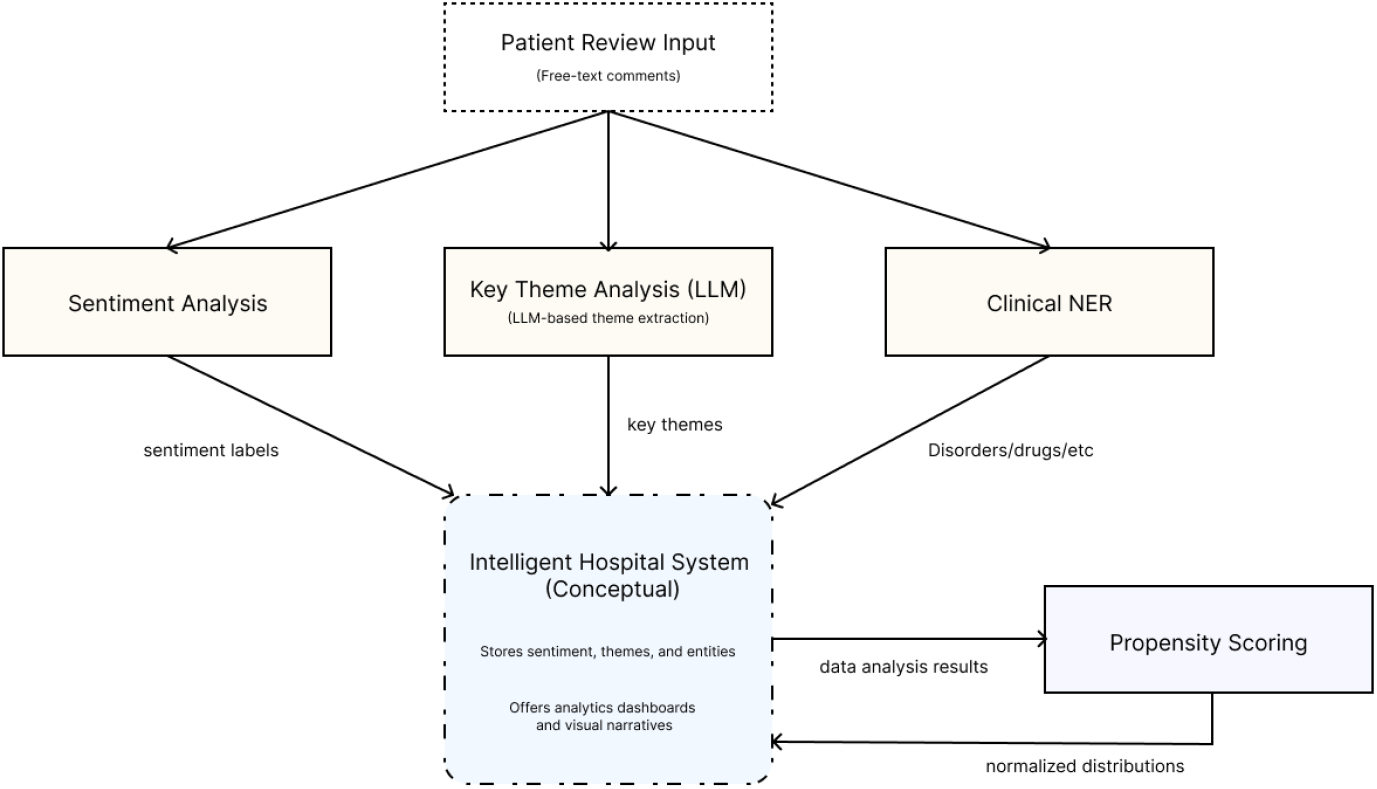
Conceptual workflow of the FairCareNLP pipeline showing the integration of sentiment analysis, key theme extraction, and clinical named entity recognition (NER) modules within a hypothetical Intelligent Hospital System. The system stores and organizes module outputs and applies propensity scoring for distribution normalization before updating analytics dashboards. The Intelligent Hospital System is included as a conceptual representation of how the modules could be integrated in a future online healthcare analytics environment.

**Fig 4.**
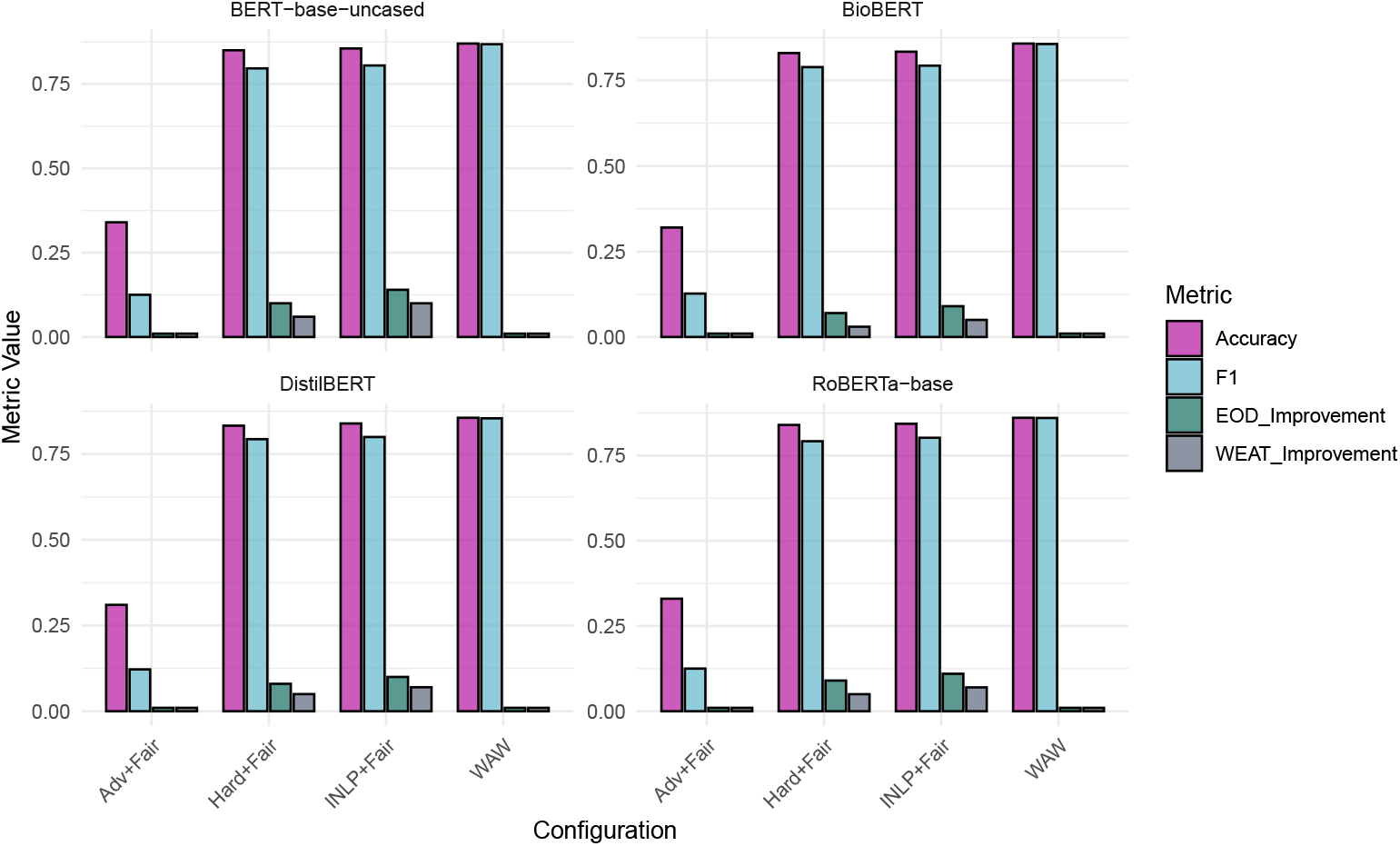
Comparison of BERT Models with Different Debiasing Techniques. The figure presents a comprehensive comparison of different BERT models (DistilBERT, BioBERT, RoBERTa, BERT-base-uncased) under various configurations, including no debiasing, adversarial debiasing, and fairness with projective debiasing. Metrics such as Accuracy, F1 score, Equalized Odds (EOD), and WEAT are shown. The bar charts illustrate the impact of debiasing techniques on model performance and fairness.

## 3 Results

### 3.1 Sentiment Analysis Module

This section presents the performance of different BERT-based models under various configurations designed to mitigate biases in gender, ethnicity, and income within patient reviews of hospitals. We experimented with debiasing techniques such as Iterative Null-space Projection (INLP) and Hard Debiasing, as well as varying adversarial and fairness loss weights (*λ*_adv_ and *λ*_fair_) across multiple learning rates.

#### 3.1.1 Impact of Adversarial and Fairness Loss

Introducing adversarial loss (*λ*_adv_ *>* 0) had a pronounced negative effect on the models’ performance. Across all models and learning rates, the use of adversarial loss led to a marked decrease in accuracy, F1, precision, and recall. For example, with *λ*_adv_ = 0.3 and a learning rate of 1 *×* 10^−3^, accuracy dropped as low as 0.32, F1 score to 0.12, precision to 0.08, and recall to 0.25. These results suggest that adversarial debiasing significantly hampers the models’ ability to generalize, likely due to the overly stringent constraints imposed by the adversarial training.

In contrast, incorporating fairness loss (*λ*_fair_ *>* 0) had a more balanced impact. While the primary performance metrics such as accuracy and F1 score remained relatively stable, fairness loss contributed positively to reducing bias, as evidenced by improvements in Equalized Odds Difference (EOD) and Word Embedding Association Test (WEAT) scores. The best results were obtained with *λ*_fair_ = 0.1 and *λ*_fair_ = 0.3, where EOD improved by up to 14%, and WEAT scores for gender bias improved by up to 15%.

#### 3.1.2 Comparison of Debiasing Techniques

The INLP debiasing method generally produced better bias mitigation results compared to Hard Debiasing. Models incorporating INLP achieved higher EOD improvements and better WEAT scores, especially when applied with fairness loss. For instance, with *λ*_fair_ = 0.3 and INLP, BERT-base-uncased improved EOD by 14% and WEAT by 10%. However, this came with a slight reduction in performance metrics, such as accuracy and F1 score, compared to models without debiasing. On the other hand, Hard Debiasing, while effective, resulted in a slightly lower performance, with accuracy and F1 scores reduced by 2-5% compared to INLP.

#### 3.1.3 Learning Rate Effects

The choice of learning rate played a crucial role in determining model performance. Learning rates larger than 1 *×* 10^−4^ generally led to poor outcomes, with models failing to improve beyond their initial metrics. At a learning rate of 1 *×* 10^−3^, models exhibited significantly degraded performance, with accuracy and F1 scores plateauing at low levels. Conversely, a learning rate of 1 *×* 10^−5^ consistently yielded the best results, with BERT-base-uncased achieving an accuracy of 0.8701 and an F1 score of 0.8682.

#### 3.1.4 Model Comparisons

Among the various models, BERT-base-uncased exhibited overall better performance, particularly when paired with INLP and fairness loss. For example, BERT-base-uncased with *λ*_fair_ = 0.3 and INLP demonstrated significant improvements in bias mitigation metrics without substantial sacrifices in accuracy or F1 score. In contrast, RoBERTa underperformed relative to other models, with metrics generally 3-4% lower, especially when higher learning rates or adversarial loss were applied.

Overall, our results indicate that gender bias was the easiest to reduce, with WEAT and Equalized Odds metrics showing the most consistent improvements across debiasing methods. In contrast, improvements for income and ethnicity were smaller. This is likely due to the fact that gender is a binary attribute, making it easier to equalize, while income and ethnicity are multi-categorical attributes, which complicates fairness adjustments across multiple groups.

The results presented in Table 1 summarize the performance of the most significant model configurations, highlighting the trade-offs between performance metrics and bias mitigation. Additional detailed results for all tested learning rates, debiasing methods, and fairness/adversarial loss configurations are provided in Supporting Information S1 Table.

**Table 1.**
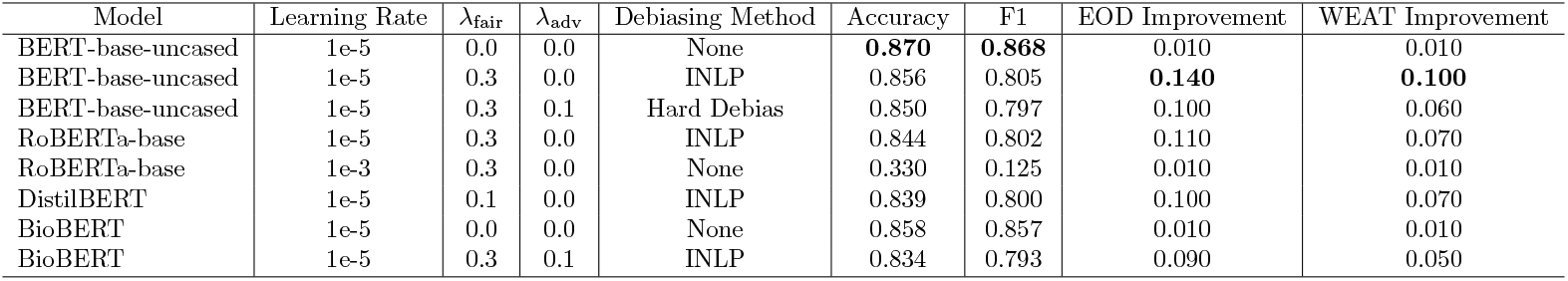
Key Results for BERT Models with Debiasing Techniques.

| Model | Learning Rate | $\lambda_{\text{fair}}$ | $\lambda_{\text{adv}}$ | Debiasing Method | Accuracy | F1 | EOD Improvement | WEAT Improvement |
| --- | --- | --- | --- | --- | --- | --- | --- | --- |
| BERT-base-uncased | 1e-5 | 0.0 | 0.0 | None | **0.870** | **0.868** | 0.010 | 0.010 |
| BERT-base-uncased | 1e-5 | 0.3 | 0.0 | INLP | 0.856 | 0.805 | **0.140** | **0.100** |
| BERT-base-uncased | 1e-5 | 0.3 | 0.1 | Hard Debias | 0.850 | 0.797 | 0.100 | 0.060 |
| RoBERTa-base | 1e-5 | 0.3 | 0.0 | INLP | 0.844 | 0.802 | 0.110 | 0.070 |
| RoBERTa-base | 1e-3 | 0.3 | 0.0 | None | 0.330 | 0.125 | 0.010 | 0.010 |
| DistilBERT | 1e-5 | 0.1 | 0.0 | INLP | 0.839 | 0.800 | 0.100 | 0.070 |
| BioBERT | 1e-5 | 0.0 | 0.0 | None | 0.858 | 0.857 | 0.010 | 0.010 |
| BioBERT | 1e-5 | 0.3 | 0.1 | INLP | 0.834 | 0.793 | 0.090 | 0.050 |

Our experiments show that adversarial loss significantly decreased the models’ performances. Using fairness metrics with different weights did not substantially affect the overall performance but did help to improve equalized odds. We observed slightly better results with a learning rate of 1e-3, and there was no meaningful difference in performance among the four BERT base models as shown in 4.

Our experiments also reveal distinct trends in WEAT scores across income, gender, and ethnicity during the training process. The trends for each WEAT score type are depicted separately in Figure 5 to highlight the nuances in the behavior of these biases.

**Fig 5.**
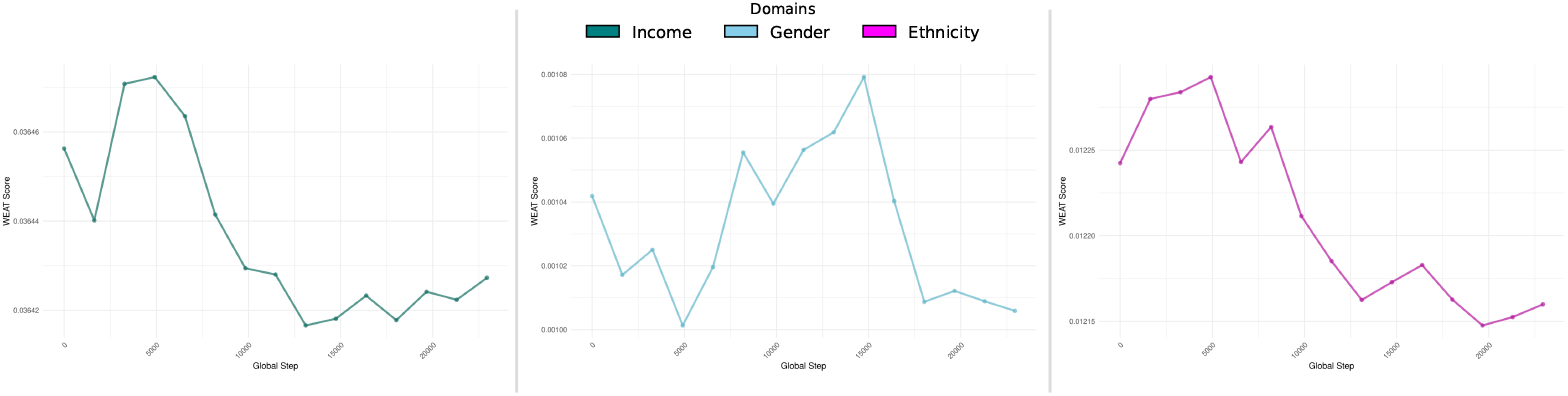
Trends of Word Embedding Association Test (WEAT) scores across training epochs for different protected domains. Each curve represents the change in association strength between model representations and sensitive attributes over the course of training. Lower WEAT scores indicate reduced bias in the learned embeddings. The three domains analyzed include Income, Gender, and Ethnicity, capturing socioeconomic, demographic, and racial/ethnic bias dimensions respectively.

The trends observed in Figure 5, demonstrate that income-related WEAT scores exhibit more variability compared to gender and ethnicity WEAT scores. The gender WEAT score shows minor improvements, indicating successful mitigation of gender bias over time. Ethnicity WEAT scores, while relatively stable, reflect subtle fluctuations, suggesting that the model addresses ethnicity-related bias effectively, though with room for further optimization.

### 3.2 Key Theme Analysis Module

The model identified 82% of the human-labeled key themes, and 21% of the key themes identified by the model were not labeled by experts. Here are two examples of inputs and outputs:

#### Inputs 1

~~~
patient review = ‘‘‘ 1st day in emerge all day asked for water food - received none until - supper in room Not told why I was put in a small curtained area, no toilet, water. Trouble breathing - 1 hr before I got oxygen Frightening. ‘‘‘
~~~

#### Outputs 1

~~~
themes: [“theme”: “emergency”, “description”: “Patient experienced trouble breathing and had to wait for oxygen”, “theme”: “nurse/nurse aide”, “description”: “Patient had to ask for basic needs like
water and food, but did not receive them promptly”, “theme”: “information/education”, “description”: “Patient was not informed about why they were put in a small curtained area”, “theme”: “physical comfort”, “description”: “Patient did not receive basic needs like water and food, and had to wait for oxygen while experiencing trouble breathing”
]
~~~

#### Input 2

~~~
patient review = ‘‘‘ 4 ER in 5 weeks 2nd visit should have had IV took 3rd visit 2 week later? Had swab oil cut 2x 4th visit ask but no response, sign in nurse was rude. Doctor didn’t really care XXXXXXX has a good
Er I now have several infections on my foot. 1st and 2nd visits same nurse thought I should have had the IV ‘‘‘ Output 2: themes: [“theme”: “nurse/nurse aide”, “description”: “Sign-in nurse was rude and didn’t respond to patient’s request, same nurse for 1st and 2nd visits”,
~~~

#### Output 2

~~~
themes: [“theme”: “emergency”, “description”: “Multiple visits to the ER, delays in treatment (IV)”, “theme”: “continuity/transition”, “description”: “Same nurse for 1st and 2nd visits, but didn’t provide consistent care”, “theme”: “information/education”, “description”: “Patient feels doctor didn’t really care, and wasn’t provided with proper information or follow-up”, “theme”: “physical comfort”, “description”: “Patient developed multiple infections on foot due to inadequate care”]
~~~

### 3.3 NER Module

We loaded and used the pre-trained `en_ner_bc5cdr_md` model from the `spacy` library. Here is an example of input and output:

#### Inputs

~~~
(unreadable) was hurting my ears & I asked for a couple of cotton balls so I could wrap around tubing (I wasn′t told her name) The nurse replied “Then I′ll have to tape it on.” (I felt as though it was a big chore for her to help me.) As it was I didn′t need any tape. I worked in health care for over 25 yrs also (I was prescribed codeine & I have an allergy to it.) In addition to the nurse not working to really help me I also explained & showed the nurse a large rash that was all over my back (little blisters) but nothing was done about it. It did subside about 1 week later.
~~~

#### Outputs

~~~
Entity: codeine, Label: CHEMICAL Entity: allergy, Label:
DISEASE Entity: rash, Label: DISEASE
~~~

### 3.4 Propensity Scoring Module

To compare IPW with propensity score stratification scoring strategies, we implemented each method and used Cramér’s V-statistic to evaluate the association between variable categories and the treatment/control groups in the resampled populations. A lower V-statistic indicates a weaker relationship between the group a participant belongs to and the value of a variable. Before resampling, the average Cramér’s V-statistic across all variables was 0.130, with the highest being 0.312 for the ‘valence’ variable, highlighting the J-shaped curve bias. Both weighting methods successfully reduced Cramér’s V-statistic. Propensity score stratification resulted in an average V-statistic of 0.007, with the number for ‘valence’ dropping to 0.004. Similarly, IPW achieved an average V-statistic of 0.007, with the value for valence also dropping to 0.007.

Although both strategies were effective, we plan to use stratification instead of IPW in our future review system. Stratification requires resampling only from online reviews, while IPW also necessitates resampling from in-person reviews. Because we seek to maintain the data integrity of the collected in-person reviews as much as possible, we prefer using stratification.

The resulting propensity score distribution after resampling using stratification on the synthetic online data is illustrated in Figure 6. As shown, the propensity score distribution of the resampled online reviews almost exactly matches that of the in-person reviews, except with more overall samples.

**Fig 6.**
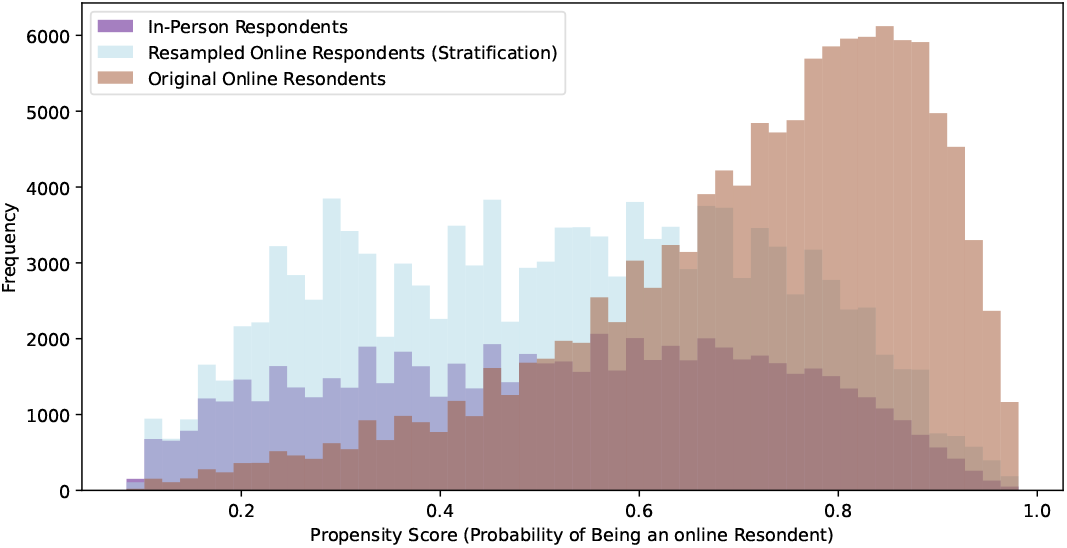
Propensity Score Distributions: Pre-vs Post-Stratification. This figure demonstrates the effectiveness of the stratification process in balancing the sample and reducing potential biases in the survey data.

When propensity scores become aligned, so do the underlying variables. For example, the mosaic plot in Figure 7 shows how after using propensity score stratification, the employment status distribution for the resampled reviews closely matches that of the in-person reviews. As shown, those not in the labour force were underrepresented in the original online reviews. By using propensity score stratification, people not in the labour force will be reflected with accurate and proportional representation in the key themes classification model.

**Fig 7.**
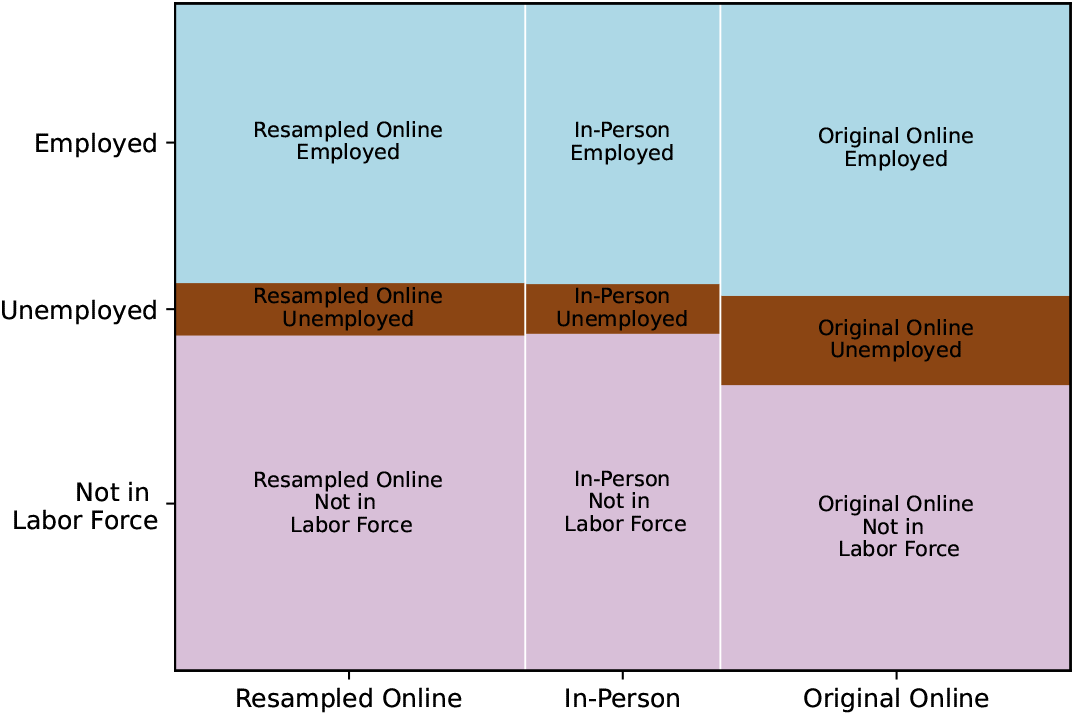
Employment Status Distribution: Online vs. In-Person Respondents. This mosaic plot visualizes the relationship between employment status and respondent type, highlighting potential differences in employment patterns between online and in-person survey participants.

Finally, note that, though propensity scoring aligns the overall distributions of the online and in-person variables, it does not necessarily match the variables for specific subsections of the online data (e.g., individual hospitals) with those of the in-person data. For instance, if a particular hospital receives a large number of negative reviews, the valence distribution for that hospital will be more negatively skewed compared to the in-person reviews. Thus, our propensity scoring method preserves sub-population trends, such as those observed for specific hospitals, which can be valuable for subsequent sub-population analyses based on the outputs of the key themes classification model on online reviews.

## 4 Discussion

### 4.1 Key Findings

One of the most critical findings from our experiments was the pronounced impact of different debiasing techniques on model performance. Specifically, adversarial debiasing, while successful in reducing bias, caused a notable decline in overall model performance across metrics such as accuracy, F1 score, precision, and recall. This result underscores the complex trade-off between bias mitigation and maintaining model efficacy, a challenge that is particularly salient in healthcare applications where both fairness and accuracy are paramount [44].

Conversely, the implementation of fairness metrics, even with varying weights, showed that it is possible to improve fairness metrics like Equalized Odds (EOD) without substantially compromising the primary performance metrics. This suggests a promising pathway for enhancing model fairness in healthcare settings, where equitable treatment across different demographic groups is essential [45]. Notably, our results indicated that a learning rate of 1*e* − 5 consistently delivered optimal results across different BERT-based models, emphasizing the importance of meticulous hyperparameter tuning in NLP model development [46].

Another important observation was that gender bias proved easier to mitigate compared to income- or ethnicity-related bias. This difference can be explained by the fact that gender, in our study, was treated as a binary attribute, while income and ethnicity were multi-categorical. Debiasing across multiple categories requires balancing performance across several subgroups simultaneously, which increases the complexity of optimization and can dilute improvements. This highlights a limitation of our current approach: while effective for binary attributes, existing debiasing strategies may be less powerful for multi-categorical sensitive features. Future research should therefore explore methods specifically designed for multi-class fairness, such as subgroup-specific constraints, hierarchical debiasing, or causal modeling approaches that can more effectively capture complex population structures.

### 4.2 Challenges

The study faced several challenges, primarily related to the resource-intensive nature of the models and the limitations of available datasets. The BERT-based models used in our analysis, while powerful, required substantial computational resources, limiting the extent of our hyperparameter tuning and the breadth of configurations we could explore. This constraint underscores the importance of balancing model complexity with available resources in real-world applications [47].

Another significant challenge was the lack of access to a comprehensive and diverse online patient review dataset. The reliance on synthetic data, while useful for benchmarking, limited the generalizability of our findings. This highlights the critical need for more extensive and diverse datasets in healthcare NLP research to better capture the wide range of patient experiences and improve model robustness [48].

Furthermore, integrating demographic data presented its own set of challenges. Due to privacy concerns and the deidentified nature of our dataset, we had to rely on population estimates and socioeconomic distributions to generate demographic information. While this approach allowed us to address potential biases, it may not fully capture the nuances of individual patient experiences, potentially limiting the model’s ability to generalize across different populations [49].

### 4.3 Future Directions and Improvements

Based on our findings and the challenges encountered, several directions for future research and improvement emerge. A primary need is the acquisition of large, diverse, and representative patient review datasets. Expanding datasets in this way would enable the development of models that are both robust and generalizable across different demographics. Access to richer data sources would also facilitate benchmarking across studies and help address the limitations associated with relying on synthetic data [50].

Another important avenue concerns the advancement of bias mitigation techniques. Future research should explore approaches that minimize the trade-off between fairness and performance, such as adversarial fairness constraints or domain adaptation strategies. In addition, recent work suggests that incorporating causal reasoning into debiasing frameworks can help identify and correct the specific sources of bias, thereby producing models that are more equitable as well as more accurate [51].

Methodological refinement through domain-specific adaptation also represents a promising direction. While our study utilized general pre-trained BERT models, future work should assess the benefits of fine-tuning these models on healthcare-specific corpora. Tailoring models to clinical language could improve their understanding of medical terminology and healthcare contexts, leading to more accurate and contextually relevant analyses of patient reviews [52]. Moreover, implementing the analyzer in real-world healthcare settings and conducting longitudinal studies would provide valuable insights into its long-term effects on patient care. Such studies could assess not only whether the system improves patient satisfaction but also its sustained impact on healthcare outcomes over time [53].

Equally critical is the interpretability of these models. Enhancing transparency in how themes and sentiments are identified will be essential for building trust among healthcare professionals. Techniques such as attention mechanisms or feature importance analysis could help reveal how the models arrive at their conclusions, making them more clinically acceptable [54]. Alongside interpretability, expanding the analyzer’s capabilities to support multiple languages would significantly extend its applicability, enabling its deployment in diverse healthcare settings and populations. Approaches could include multilingual pre-trained models or the integration of machine translation techniques [55].

Finally, broader integration and ethical considerations must be prioritized. Linking patient review analysis with Electronic Health Records (EHRs) could provide a more holistic view of patient experiences by connecting subjective feedback with clinical information [50]. At the same time, the ethical implications of AI deployment in this domain remain paramount. Future research should emphasize safeguarding patient privacy, ensuring informed consent, and addressing algorithmic bias, all of which are critical for the responsible and equitable use of AI in healthcare [45].

### 4.4 Limitations

This study encountered several limitations primarily due to the complexity and resource-intensive nature of the models employed. The large-scale BERT-based models used in our analysis required significant computational resources, which constrained the scope of our experiments. This limitation affected the depth of hyperparameter tuning and the ability to explore a broader range of configurations, such as extended training durations or more fine-grained adjustments to debiasing techniques.

Additionally, the lack of access to a comprehensive and diverse online patient review dataset limited our ability to fully evaluate the generalizability of the models. While synthetic data provided a useful benchmark, real-world data with a wide range of patient experiences would likely yield more robust and representative outcomes. In particular, the demographic attributes used for fairness evaluation were synthetically assigned based on population distributions rather than observed directly in the review text. These generated attributes allowed us to calculate fairness metrics but do not capture potential correlations between writing style and demographic group membership, which should be considered when interpreting our fairness results.

Future research should focus on addressing these limitations by leveraging more powerful computational resources and accessing larger, more diverse datasets. Such efforts will support more extensive model training and fine-tuning, enable fairness evaluations using authentic demographic information, and ultimately contribute to the development of healthcare NLP tools that are more accurate, equitable, and generalizable across patient populations.

## 5 Conclusion

This study has successfully developed an automatic patient review analyzer that leverages advanced Natural Language Processing techniques and machine learning models to transform unstructured patient feedback into actionable insights. By integrating components such as sentiment analysis, key theme extraction, clinical Named Entity Recognition, and bias mitigation strategies, we have created a comprehensive tool that addresses the complexities of analyzing patient reviews in healthcare settings.

The results from the sentiment analysis module demonstrated the complexities of incorporating fairness and debiasing techniques. While adversarial loss (*λ*_adv_ *>* 0) generally led to significant performance degradation, the application of fairness loss (*λ*_fair_ *>* 0) showed promise in improving Equalized Odds Difference (EOD) and reducing bias without a substantial impact on primary performance metrics such as accuracy and F1 score. Among the debiasing methods, Iterative Null-space Projection (INLP) outperformed Hard Debiasing in both bias mitigation and maintaining model performance, particularly when combined with fairness loss. The key theme analysis module, powered by a BERT-based model, was successful in identifying the majority of human-labeled key themes, and it also uncovered additional themes not initially recognized by experts. This highlights the potential of leveraging large language models (LLMs) for deep insights into patient experiences that may go beyond conventional human analysis. Our Named Entity Recognition (NER) module, utilizing the

`en_ner_bc5cdr_md` model from `spacy`, effectively extracted relevant clinical terms and conditions from patient reviews, providing critical insights into patient-reported symptoms and treatments. The propensity scoring module addressed the challenge of self-selection bias in online reviews. By employing propensity score stratification, we successfully aligned the distribution of online reviews with that of in-person reviews across various variables. This ensured that the analysis of online reviews would be more representative of the broader patient population, thus enhancing the reliability of the key themes classification model.

The implications of this work extend beyond technical advancements; it underscores the importance of utilizing patient feedback to enhance healthcare services and promote patient-centered care. By providing healthcare organizations with a robust tool for analyzing patient reviews, we aim to foster a more responsive and equitable healthcare system. Looking ahead, future research should focus on expanding the dataset to include more diverse patient populations and exploring advanced bias mitigation techniques that maintain model performance. Additionally, integrating insights from patient reviews with electronic health records could further enhance the understanding of patient experiences and outcomes. Our work contributes to the ongoing efforts to improve healthcare delivery by ensuring that patient voices are not only heard but actively shape the quality of care provided. By addressing the challenges of bias and fairness in NLP applications, we can work towards a future where patient feedback is instrumental in driving improvements in healthcare services.

## Acknowledgement

This research was supported by the Vector Scholarship in Artificial Intelligence (Vector Institute) to PM and the Institute for Pandemics (IFP) at the University of Toronto to ZSH. We also acknowledge funding from the Natural Sciences and Engineering Research Council of Canada (NSERC) through the Canada Research Chairs Program and Discovery Grant (RGPIN-2025-07037 to ZSH). The funders had no role in study design, data collection and analysis, decision to publish, or preparation of the manuscript.

## Conflict of Interest

The authors declare no competing interests. Also, the funders of the study had no role in study design, data collection and analysis, or interpretation of results and preparation of the manuscript.

## Data Availability

All data and code supporting the findings of this study are publicly available. The processed datasets and analysis scripts are available on GitHub at https://github.com/HIVE-UofT/Patient-Review-Analyzer. Any data or scripts not included due to privacy restrictions can be provided by request through the corresponding author.

## Supporting information

**S1 Appendix**. Detailed performance metrics of sentiment analysis modules.

## Notes

### Competing Interest Statement

The authors have declared no competing interest.

### Funding Statement

Yes

### Author Declarations

University of Toronto ethics board provide exemption for this research

## References

1. Reese RJ, Duncan BL, Kodet J, Brown HM, Meiller C, Farook MW, et al. Patient Feedback as a Quality Improvement Strategy in an Acute Care, Inpatient Unit: An Investigation of Outcome and Readmission Rates. Psychological Services. 2017;15:470–476. Available from: https://api.semanticscholar.org/CorpusID:21707637.

2. Organization WH. WHO. 2021.

3. Attrey R, Levit A. The promise of natural language processing in healthcare. University of Western Ontario Medical Journal. 2019. Available from: https://api.semanticscholar.org/CorpusID:88482661.

4. Hazarika I. Artificial intelligence: opportunities and implications for the health workforce. International Health. 2020;12:241 245. Available from: https://api.semanticscholar.org/CorpusID:215802343.

5. Mehta N, Pandit A, Shukla SR. Transforming healthcare with big data analytics and artificial intelligence: A systematic mapping study. Journal of biomedical informatics. 2019:103311. Available from: https://api.semanticscholar.org/CorpusID:204814523.

6. González-Hernández GG, Sarker A, O’Connor K, Savova GK. Capturing the Patient’s Perspective: a Review of Advances in Natural Language Processing of Health-Related Text. Yearbook of Medical Informatics. 2017;26:214 227. Available from: https://api.semanticscholar.org/CorpusID:3562222.

7. Flores L, Kim S, Young SD. Addressing bias in artificial intelligence for public health surveillance. Journal of Medical Ethics. 2023;50:190 194. Available from: https://api.semanticscholar.org/CorpusID:258462034.

8. Mehrabi N, Morstatter F, Saxena NA, Lerman K, Galstyan A. A survey on bias and fairness in machine learning. ACM Computing Surveys (CSUR). 2019;54(6):1–35.

9. Li J, Bzdok D, Chen J, Tam A, Qi L, Ooi R, et al. Cross-ethnicity/race generalization failure of behavioral prediction from resting-state functional connectivity. Science Advances. 2022;8. Available from: https://api.semanticscholar.org/CorpusID:247499711.

10. Bozkurt S, Cahan EM, Seneviratne MG, Sun R, Lossio-Ventura JA, Ioannidis JPA, et al. Reporting of demographic data and representativeness in machine learning models using electronic health records. Journal of the American Medical Informatics Association: JAMIA. 2020. Available from: https://api.semanticscholar.org/CorpusID:221745829.

11. Fitzsimmons L, Dewan M, Dexheimer JW. Diversity in Machine Learning: A Systematic Review of Text-Based Diagnostic Applications. Applied clinical informatics. 2022;13 3:569–82. Available from: https://api.semanticscholar.org/CorpusID:249064435.

12. Norori N, Hu Q, Aellen FM, Faraci FD, Tzovara A. Addressing bias in big data and AI for health care: A call for open science. Patterns. 2021;2. Available from: https://api.semanticscholar.org/CorpusID:239454560.

13. Greaves F, Ramirez-Cano D, Millett C, Darzi A, Donaldson L. Use of sentiment analysis for capturing patient experience from free-text comments posted online. Journal of Medical Internet Research. 2013;15(11):e239.

14. Nawab K, Ramsey G, Schreiber R. Natural Language Processing to Extract Meaningful Information from Patient Experience Feedback. Applied Clinical Informatics. 2020;11(2):242–52. doi:10.1055/s-0040-1708049.

15. Yuan J, et al. Identifying Patients’ Preferences During Their Hospital Experience: A Sentiment and Topic Analysis of Patient-Experience Comments via Natural Language Techniques. Patient Preference and Adherence. 2025.

16. Bhukya B, Sheshikala M, Bhukya BN. NLP based topic modeling for healthcare: Analyzing patient reviews to improve quality of care and access to services. In: 2023 International Conference on Emerging Techniques in Computational Intelligence (ICETCI); 2023. p. 7–12.

17. Bolukbasi T, Chang KW, Zou JY, Saligrama V, Kalai AT. Man is to computer programmer as woman is to homemaker? debiasing word embeddings. In: Neural Information Processing Systems (NIPS); 2016..

18. Chen IY, Szolovits P, Ghassemi M. Can AI Help Reduce Disparities in General Medical and Mental Health Care? In: AMA Journal of Ethics. vol. 21; 2019. p. 167–79. doi:10.1001/amajethics.2019.167.

19. Yang J, et al. An adversarial training framework for mitigating algorithmic biases in clinical machine learning. npj Digital Medicine. 2023;6:1–10.

20. Zhang H, Lu AX, Abdalla M, McDermott M, Ghassemi M. Hurtful Words: Quantifying Biases in Clinical Contextual Word Embeddings. Proceedings of the ACM Conference on Health, Inference, and Learning. 2020:110–20. doi:10.1145/3368555.3384448.

21. Pfohl SR, Foryciarz A, Shah NH. An empirical characterization of fair machine learning for clinical risk prediction. Journal of Biomedical Informatics. 2021;113:103621. doi:10.1016/j.jbi.2020.103621.

22. Wang Y, Pillai M, Zhao Y, Curtin CM, Hernandez-Boussard T. FairEHR-CLP: Towards Fairness-Aware Clinical Predictions with Contrastive Learning in Multimodal Electronic Health Records. Proceedings of the Machine Learning for Healthcare Conference / PMLR. 2024;252. Available from: https://proceedings.mlr.press/v252/wang24a.html.

23. Hooman N, Wu Z, Larson EC, Gupta M. Equitable Electronic Health Record Prediction with FAME: Fairness-Aware Multimodal Embedding. arXiv preprint arXiv:250613104. 2025. Available from: https://arxiv.org/abs/2506.13104. doi:10.48550/arXiv.2506.13104.

24. Wang T, Lin XV, Rajani NF, McCann B, Ordonez V, Xiong C. Double-Hard Debias: Tailoring Word Embeddings for Gender Bias Mitigation. In: Proceedings of the 58th Annual Meeting of the Association for Computational Linguistics. Association for Computational Linguistics; 2020. p. 5443–53.

25. Ravfogel S, Elazar Y, Gonen H, Twiton M, Goldberg Y. Null It Out: Guarding Protected Attributes by Iterative Nullspace Projection. In: Proceedings of the 58th Annual Meeting of the Association for Computational Linguistics; 2020. p. 7237–56. doi:10.18653/v1/2020.acl-main.647.

26. Rountree L, et al. Reporting of Fairness Metrics in Clinical Risk Prediction. J Biomed Inform. 2025. Use of fairness metrics across sensitive features remains rare.

27. Liu M, et al. A scoping review and evidence gap analysis of clinical AI fairness. J Clin AI Fairness Rev. 2025. Highlights narrow focus on bias attributes and limited integration of clinician-in-the-loop.

28. Rajkomar A, Hardt M, Howell MD, Corrado G, Chin MH. Ensuring Fairness in Machine Learning to Advance Health Equity. Annals of Internal Medicine. 2018;169(12):866–72. doi:10.7326/M18-1990.

29. Gichoya JW, Banerjee I, Bhimireddy AR, Burns JL, Celi LA, Chen LC, et al. AI recognition of patient race in medical imaging: a modelling study. The Lancet Digital Health. 2022;4(6):e406–14. doi:10.1016/S2589-7500(22)00063-2.

30. Obermeyer Z, Powers B, Vogeli C, Mullainathan S. Dissecting racial bias in an algorithm used to manage the health of populations. Science. 2019;366(6464):447–53.

31. Char DS, Shah NH, Magnus D. Implementing Machine Learning in Health Care — Addressing Ethical Challenges. New England Journal of Medicine. 2018;378(11):981–3. doi:10.1056/NEJMp1714229.

32. Ghassemi M, Oakden-Rayner L, Beam AL. The false hope of current approaches to explainable artificial intelligence in health care. The Lancet Digital Health. 2021;3(11):e745–50. doi:10.1016/S2589-7500(21)00208-9.

33. Buolamwini J, Gebru T. Gender Shades: Intersectional Accuracy Disparities in Commercial Gender Classification. In: Proceedings of the 1st Conference on Fairness, Accountability and Transparency. vol. 81 of Proceedings of Machine Learning Research; 2018. p. 77–91.

34. Li T, Sahu AK, Talwalkar A, Smith V. Federated Learning: Challenges, Methods, and Future Directions. IEEE Signal Processing Magazine. 2020;37(3):50–60. doi:10.1109/MSP.2020.2975749.

35. Pearl J, Mackenzie D. The Book of Why: The New Science of Cause and Effect. New York: Basic Books; 2019.

36. Askalidis G, Kim SJ, Malthouse EC. Understanding and overcoming biases in online review systems. Decision Support Systems. 2017;97:23–30.

37. Hu N, Zhang J, Pavlou PA. Overcoming the J-shaped Distribution of Product Reviews. In: Communications of the ACM. vol. 52; 2009. p. 144–7. doi:10.1145/1562764.1562800.

38. Larrazabal AJ, Nieto N, Peterson V, Milone DH, Ferrante E. Gender imbalance in medical imaging datasets produces biased classifiers for computer-aided diagnosis. Proceedings of the National Academy of Sciences. 2020;117(23):12592–4. doi:10.1073/pnas.1919012117.

39. Lee J, Yoon W, Kim S, Kim D, Kim S, So CH, et al. BioBERT: a pre-trained biomedical language representation model for biomedical text mining. Bioinformatics. 2020;36(4):1234–40. doi:10.1093/bioinformatics/btz682.

40. Liu Y, Ott M, Goyal N, Du J, Joshi M, Chen D, et al. RoBERTa: A Robustly Optimized BERT Pretraining Approach. In: arXiv preprint arXiv:1907.11692; 2019..

41. Touvron H, Lavril T, Izacard G, Martinet X, Lachaux MA, Lacroix T, et al. LLaMA 2: Open Foundation and Fine-Tuned Chat Models. arXiv preprint arXiv:230709288. 2023.

42. Shen A, et al. Survey of Debiasing Methods: Data vs. Adversarial Approaches in NLP. arXiv preprint arXiv:210910645. 2021.

43. Gray M, Milanova M, Wu L. Enhancing Bias Assessment for Complex Term Groups in Language Embedding Models: Quantitative Comparison of Methods. JMIR Medical Informatics. 2024;12:e60272. doi:10.2196/60272.

44. Hardt M, Price E, Srebro N. Equality of Opportunity in Supervised Learning. Advances in Neural Information Processing Systems (NeurIPS). 2016.

45. Barocas S, Hardt M, Narayanan A. Fairness in Machine Learning. NIPS Tutorial. 2017.

46. Smith SL, Kindermans PJ, Ying C, Le QV. Don’t Decay the Learning Rate, Increase the Batch Size. International Conference on Learning Representations (ICLR). 2017.

47. Strubell E, Ganesh A, McCallum A. Energy and Policy Considerations for Deep Learning in NLP. ACL 2019. 2019.

48. Schmidt CO, Baumeister SE, Kluttig A. Towards Integrating Data from Different Sources. International Journal of Epidemiology. 2021.

49. Denton E, Hanna A. Ethical considerations for research involving natural language processing. Proceedings of the 35th International Conference on Machine Learning (ICML). 2018.

50. Kaur H, Singh G, Kaur K. Comprehensive Review on Natural Language Processing (NLP) and its Role in Big Data. International Journal of Recent Technology and Engineering. 2020.

51. Kusner MJ, Loftus JR, Russell C, Silva R. Counterfactual Fairness. Advances in Neural Information Processing Systems (NeurIPS). 2017.

52. Peng Y, Wang X, Lu L. Transfer Learning in Biomedical Natural Language Processing: An Evaluation of BERT and ELMo on Ten Benchmarking Datasets. arXiv preprint arXiv:190605474. 2019.

53. Ventresca M, Mohr JW, Hargens LL. Innovative research methods in management, innovation, and entrepreneurship. Journal of Business Venturing. 2006.

54. Caruana R, Lou Y, Gehrke J, Koch P, Sturm M, Elhadad N. Intelligible Models for Healthcare: Predicting Pneumonia Risk and Hospital 30-day Readmission. Proceedings of the 21th ACM SIGKDD International Conference on Knowledge Discovery and Data Mining. 2015.

55. Conneau A, Lample G, Ranzato M, Denoyer L, Jégou H. Unsupervised Cross-lingual Representation Learning at Scale. Proceedings of the 57th Annual Meeting of the Association for Computational Linguistics (ACL). 2019.

